# A Service Evaluation of the Active Together Multi-Modal Cancer Prehabilitation and Rehabilitation Service

**DOI:** 10.1101/2025.08.20.25334050

**Authors:** Nik Kudiersky, Kerry Rosenthal, Gabriella Frith, Gail Phillips, Carol Keen, Shaun Barratt, Diana M Greenfield, Gary Mills, Anna Myers, Robert Copeland, Liam Humphreys

## Abstract

**Background:** Active Together is a clinician-led cancer prehabilitation and rehabilitation service for adults with colorectal, lung, or upper gastrointestinal (GI) cancer receiving curative treatment in Sheffield, UK. This service evaluation assessed the feasibility, engagement, and impact of a multimodal intervention—combining exercise, dietetic, and psychological support— delivered across four phases: prehabilitation (pre-treatment), maintenance rehabilitation (during treatment), restorative rehabilitation (up to 12 weeks post-treatment), and supportive rehabilitation (12 to 24 weeks post-treatment).

**Methods:** Between March 2022 and May 2024, 847 patients were referred; 685 enrolled (80.9%), and 649 consented to evaluation (mean age 68). Outcomes included six-minute walk test (6MWT), sit-to-stand (30s/60s), handgrip strength, EQ-5D-5L index and visual analogue scale (VAS), FACIT-Fatigue, PHQ-9, and GAD-7. Wilcoxon signed-rank tests were used to assess change scores across each service phase.

**Results:** Ninety-three percent completed prehabilitation, and 62% completed the full programme (median duration 44 weeks). After prehabilitation, improvements were observed for 6MWT (+27m, p<0.001), 30s sit-to-stand (+2 reps, p<0.01), EQ-5D VAS (+7.5 points, p<0.01), fatigue (+2.5 points, p<0.01), and PHQ-9 scores (−1 point, p<0.01). Following rehabilitation, 6MWT improved by +20m above baseline, EQ-5D VAS increased +7.5–10 points, and PHQ-9 and GAD-7 scores reduced by ≈2 points (all p<0.01). Recovery trajectories differed by sex and cancer type, with males and individuals with upper GI cancer experiencing greater post-treatment decline and less complete recovery.

**Conclusion:** Active Together demonstrated high engagement and clinically meaningful improvements in cardiorespiratory fitness, and quality of life. While the absence of a control group limits conclusions about effectiveness, the findings suggest that multimodal prehabilitation and rehabilitation in routine cancer care is feasible, supports functional recovery, and improves wellbeing.

## Introduction

Multi-modal cancer prehabilitation and rehabilitation are complex interventions designed to optimise cancer-related health outcomes across the cancer care continuum (1). Intervention components typically comprise exercise, nutrition, and / or psychological support delivered by healthcare professionals and / or exercise professionals with specialist cancer training (1). Emerging evidence from randomised controlled trials indicates that uni-modal (typically exercise only) and multi-modal (exercise plus nutrition and / or psychology support) interventions are beneficial for cancer treatment outcomes (1). Key benefits include shorter hospital stays (2), enhanced physical function (1), improved body composition (3), decreased fatigue (4), improved psychological wellbeing, and an enhanced quality of life (5).

The decision to commission cancer prehabilitation and rehabilitation services requires robust evidence that these interventions work in real-world settings, beyond controlled research environments (6). However, translating research findings into clinical practice is challenging (7–9). Clinical trials often have relatively small sample sizes, strict inclusion criteria, highly standardised interventions, and are susceptible to healthy user bias. In contrast, real-world clinical services often manage high caseloads of patients with complex multimorbidity, including people living in economically disadvantaged regions who may face more barriers to accessing care (10). Real-world cancer prehabilitation and rehabilitation services are currently being trialled through various models, including services delivered in healthcare settings or in collaboration with leisure sector providers.

### Active Together Cancer Prehabilitation and Rehabilitation Service

Active Together is a cancer prehabilitation and rehabilitation service operating in South Yorkshire–a region in Northern England with a population of 1.4 million (11). Over a third of South Yorkshire’s residents live in areas of high deprivation, and the region as a whole faces a higher cancer incidence, lower life expectancy, and greater burden of chronic health conditions compared to national averages (11). Active Together is a clinician-led multi-modal service comprising exercise specialists (Level 4), physiotherapists, a dietitian, and a psychologist. All components of the service incorporate behaviour change techniques, guided by a “What matters to you?”, patient-centred approach (12). The first phase of the Active Together service, delivered in Sheffield from March 2022 to May 2024, accepted referrals for adults with a primary diagnosis of lung, colorectal or upper gastrointestinal (GI) cancer, with scheduled curative treatment in Sheffield. The service consists of four phases spanning the cancer care continuum (Figure 1), including prehabilitation (before treatment), maintenance (during treatment), restorative (up to three months after treatment) and supportive (up to six months after treatment) rehabilitation.

**Figure 1.**
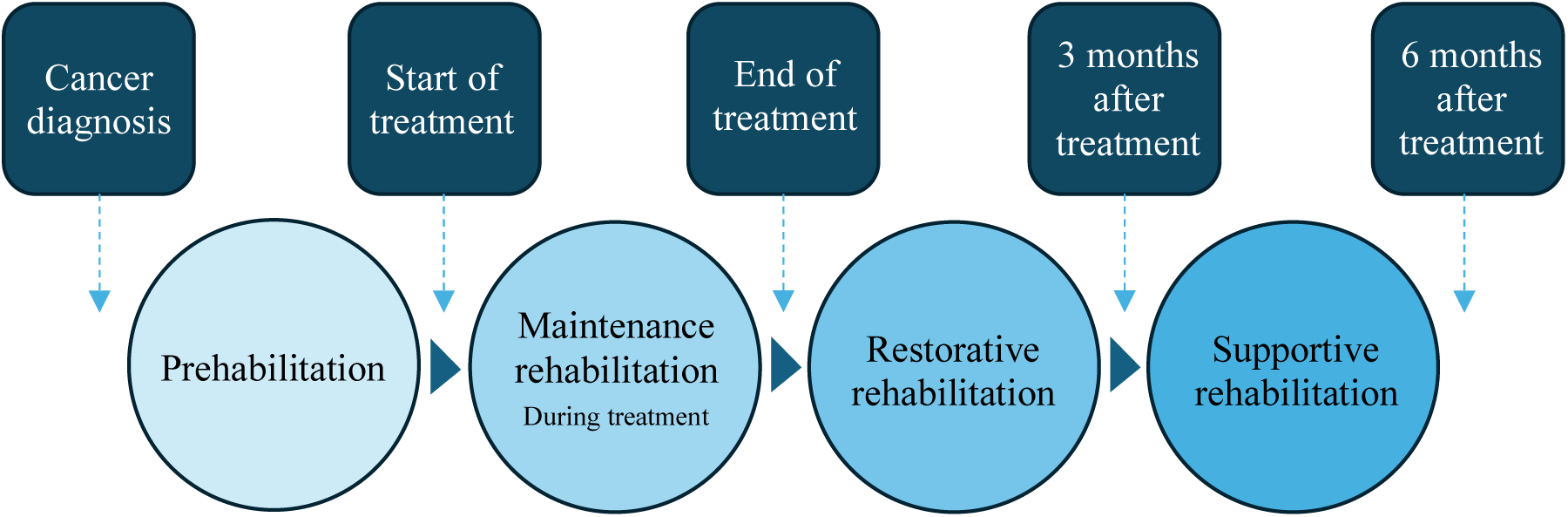
Active Together service pathway.

Using a combination of patient reported outcome measures and clinical expertise, Active Together patients are categorised as Specialist (high complexity), Targeted (moderate complexity) or Universal (low complexity) for exercise, nutritional, and psychological support. For example, a Specialist patient may present with very low functional capacity alongside poorly managed cardiac or metabolic conditions, or significant mental health concerns. In contrast, a Universal patient may demonstrate normal fitness levels, well-controlled comorbidities, and average mood scores. Tailored support is then offered, appropriate to patients’ needs. Healthcare professionals referring patients to Active Together are encouraged to avoid excluding individuals based on assumptions about their ability to engage in exercise. A detailed overview of the Active Together service design has been published (13). An ongoing service evaluation, conducted by Sheffield Hallam University, is embedded within Active Together to understand the real-world impact of the service. Here, measures of uptake, engagement, physical function, fatigue and psychological wellbeing are presented from the first phase of the service.

## Methods

### Study Design and Setting

A service evaluation of the Active Together cancer prehabilitation and rehabilitation service delivered in Sheffield, United Kingdom. This evaluation utilised a pre-post intervention design with no control group. Data were collected between March 2022 and May 2024 as part of routine clinical practice. The protocol for this service evaluation has been published elsewhere (14).

### Participants

Inclusion criteria: 1) medically stable, 2) diagnosis of colorectal, lung, or upper gastrointestinal (GI) cancer, 3) scheduled for curative treatment in Sheffield, 4) cognitively able to engage in the service. All patients who completed at least two assessments were included in the service evaluation analysis. During the initial assessment, patients were asked to provide verbal or written consent for their data to be included in the service evaluation. Patients who declined were excluded from analysis. Participant characteristics are summarised in Table 1.

**Table 1.** Patient characteristics.

| Cancer Type | Phase | Sample size (n) | Age (Mean ± SD) | % Female | % White | IMD (Mean ± SD) |
| --- | --- | --- | --- | --- | --- | --- |
| Colorectal | Prehabilitation | 91 | 70 ± 9 | 38% | 98% | 5 ± 3 |
|  | Maintenance | 141 | 69 ± 10 | 39% | 96% | 6 ± 3 |
|  | Restorative | 88 | 69 ± 10 | 35% | 96% | 6 ± 3 |
|  | Supportive | 83 | 71 ± 9 | 41% | 99% | 6 ± 3 |
| Lung | Prehabilitation | 40 | 69 ± 9 | 58% | 94% | 4 ± 3 |
|  | Maintenance | 84 | 67 ± 11 | 49% | 93% | 4 ± 3 |
|  | Restorative | 40 | 68 ± 12 | 50% | 91% | 4 ± 3 |
|  | Supportive | 45 | 70 ± 11 | 44% | 95% | 4 ± 3 |
| Gastro-intestinal | Prehabilitation | 28 | 68 ± 7 | 25% | 96% | 5 ± 3 |
|  | Maintenance | 60 | 65 ± 9 | 23% | 97% | 4 ± 3 |
|  | Restorative | 31 | 65 ± 8 | 23% | 94% | 4 ± 3 |
|  | Supportive | 23 | 65 ± 10 | 22% | 100% | 4 ± 3 |
| Total | Prehabilitation | 159 | 69 ± 9 | 41% | 97% | 5 ± 3 |
|  | Maintenance | 285 | 68 ± 10 | 39% | 95% | 5 ± 3 |
|  | Restorative | 159 | 68 ± 10 | 36% | 95% | 5 ± 3 |
|  | Supportive | 151 | 70 ± 10 | 39% | 98% | 5 ± 3 |

IMD, Index of Multiple Deprivation (21). Female = biological female

### Intervention

The Active Together service has been described in detail previously (14). A brief overview of each component of the service is provided below.

### Exercise Component

The prehabilitation phase focused primarily on increasing cardiorespiratory fitness to enhance resilience to cancer treatment. The duration of the prehabilitation phase varied based on treatment waiting periods. The maintenance phase focused on engaging in manageable physical activities during treatment and minimising sedentary behaviour where possible, with the aim of mitigating functional decline. The duration of this phase also varied depending on treatment duration. The restorative and supportive rehabilitation phases—each lasting up to 12 weeks —aimed to restore cardiorespiratory fitness and skeletal muscle function back to pre-treatment levels, while empowering behaviour change for long-term sustained engagement in regular exercise. Patients across all risk levels were encouraged to meet the UK Chief Medical Officer’s Physical Activity Guidelines (15): 150 minutes of moderate-intensity activity per week or 75 minutes of vigorous-intensity activity (where appropriate), plus muscle-strengthening and balance exercises on at least two days, and to reduce sedentary activity. Patients were typically offered one supervised exercise session per week (in-person or virtual), and were encouraged to complete further unsupervised exercise sessions in their preferred setting, facilitated with a printed exercise programme and online videos. Supervised exercise sessions were delivered via a circuit training format using a variety of fitness equipment (e.g., steps, ergometers, dumbbells, resistance bands), and patients were encouraged to use affordable exercise equipment for home-based sessions.

### Dietetic Component

All patients received healthy eating educational resources and were invited to attend an in-person nutritional workshop, delivered by a dietitian. Patients classified as Specialist (higher risk) were offered one-to-one dietetic support. This cohort included individuals with conditions such as dysphagia, poorly controlled diabetes, gastrointestinal disorders, recent rapid weight loss, or people managing a stoma. Dietetic interventions focused on optimising energy and protein intake, incorporating oral nutritional supplements where appropriate, alongside symptom management strategies.

### Psychological Component

All patient-facing Active Together staff completed Level 2 psychological skills training, enabling them to identify psychological distress and apply techniques such as problem solving (16). Patients received digital and printed resources to help manage anxiety and depression. Additionally, all delivery staff were trained in motivational interviewing, equipping them with techniques to support and empower patients in making positive lifestyle changes. Patients with more complex needs, such as high levels of anxiety or low mood, were offered a targeted intervention from a clinical psychologist.

### Fatigue Management

Support was provided to address both the psychological and physical aspects of cancer-related fatigue. This included the abovementioned psychological interventions, as well as education on pacing techniques to help patients conserve energy.

### Outcome Measures

Physical function was assessed using measures of cardiorespiratory fitness (six-minute walk test), upper limb strength (handgrip strength), lower limb muscular strength (30-second sit-to-stand test), and lower limb strength-endurance (60-second sit-to-stand test). Psychological wellbeing was evaluated via measures of quality of life (EuroQol Five-Dimension Five-Level; EQ-5D-5L) (17), depression (9-item Patient Health Questionnaire; PHQ-9) (18), anxiety (7-item Generalised Anxiety Disorder; GAD-7) (19). Fatigue was assessed using the Functional Assessment of Chronic Illness Therapy-Fatigue (FACIT-Fatigue) (20). All patient-facing staff received training in standardised data collection techniques. Uptake and engagement were assessed using referral and attendance data plus patient feedback forms. All patient assessment data were recorded in clinical records as part of usual care and exported for analysis by the service evaluation team in a pseudonymised format.

### Data Collection Timepoints

Data were collected as part of routine clinical assessments at the following timepoints:

1. Before starting prehabilitation phase
2. End of prehabilitation phase (pre-treatment)
3. End of maintenance rehabilitation phase (post-treatment)
4. End of restorative rehabilitation phase
5. End of supportive rehabilitation phase

### Statistical Analysis

The primary analysis evaluated changes in outcome measures across each phase of the Active Together service for all patients (tumour groups and sexes combined) who had a baseline assessment and at least one other assessment. Secondary analyses were performed to determine if results differed by sex or cancer type, with sex defined as biological male or female. Outcome measure data were not normally distributed; change scores were reported as pseudomedians with 95% confidence intervals. Analytical statistics involved pairwise Wilcoxon signed-rank tests to compare end-of-phase scores with baseline values, presented as change scores. Cohen’s d effect sizes were calculated to quantify the magnitude of change for overall group-level comparisons and are reported in the appendix. Benjamini-Hochberg corrections were applied to control for multiple comparisons. Engagement data were summarised using descriptive statistics. A p-value of less than 0.05 was considered statistically significant. Effect sizes (Cohen’s *d*) were reported for overall group comparisons only, using conventional thresholds (0.2 = small, 0.5 = medium, 0.8 = large). All statistical analyses were performed using R (version 4.2.0). All analyses were conducted by academics employed by Sheffield Hallam University, funded by Yorkshire Cancer Research. Data were collected between March 2022 and May 2024.

### Ethical Considerations

As this service evaluation used routinely collected clinical data and there was no change in patient care, formal ethical approval was not required per local governance policies. Data were pseudonymised prior to analysis to ensure patient confidentiality. This service evaluation was approved by the Sheffield Teaching Hospitals NHS Foundation Trust Clinical Effectiveness Unit (Reference: 11115-May 19, 2022).

### Results

#### Uptake

Between March 2022 and May 2024, Active Together received 847 referrals for patients with colorectal, lung, and upper gastrointestinal (GI) cancer. One-hundred-and-sixty-two patients declined the service, resulting in an acceptance rate of 80.9%. Acceptance rates were 86.2%, 85.8% and 72.7% for upper GI, colorectal, and lung cancer cohorts, respectively. Thirty-six patients opted out of NHS Data Sharing, leaving 649 patient datasets for analysis. Group-level patient characteristics are displayed in Table 1.

#### Intervention Engagement

The prehabilitation phase had a completion rate of 93%, with a median duration of five weeks. Engagement gradually declined across rehabilitation phases; the main reasons for attrition were: self-managing (34.3%); no contact from patient (17.1%); too unwell (16.6%); using another service (13.7%). Eighty percent completed up to the maintenance phase (median duration = 10 weeks), 66% completed up to the restorative phase (median duration = 15 weeks), and 62% completed all phases of the service up to end of the supportive phase (median duration = 14 weeks) over a total median duration of 44 weeks.

#### Intervention Characteristics

Data on exercise dose characteristics, such as average exercise intensity per session, rate of progression, and the total number of exercise sessions per week were not routinely recorded. Attendance data were also unavailable for analysis. A separate (unpublished) clinical audit determined that the exercise intensity for supervised sessions was typically 40–50% heart rate reserve (moderate intensity), with rating of perceived exertion scores generally ranging from 11-13 out of 20 (light to somewhat hard). The content of one-to-one dietitian and psychologist interventions were not audited as these sessions were tailored to patients’ individual needs and goals. Consequently, the “dose” of these interventions remains undefined.

### Physical Function Outcomes

#### Cardiorespiratory Fitness

The median baseline 6MWT distance was 487m for male and 411m for female patients. These scores are lower than normative values for community dwelling older adults (aged 60-80 years) (22), and similar lung cancer patients (median age 69 years, IQR 63–74). No between-group differences (sex or tumour group) were identified, therefore pooled change scores are reported. Figure 2 displays changes in 6MWT distance. Patients’ walking distance increased after the prehabilitation phase, with a median score of +27m (p<0.001, Cohen’s d=0.99). 6MWT distance declined after treatment compared to baseline (−17m, p<0.001, Cohen’s d=0.59). This reduction was less than that observed in clinical trial control groups who do not engage in cancer prehabilitation (−28m, measured 4-9 weeks post-surgery) (23,24), indicating that the expected post-treatment deconditioning may have been mitigated. Following the post-treatment restorative and supportive rehabilitation phases, patients’ walking distance increased, with a final median improvement of 20m above baseline (both phases, p<0.01, Cohen’s d>0.5), and a median 37m increase above the post-treatment score. The minimum clinically important difference (MCID) for 6MWT is 14-42 metres for older adults with chronic disease, including lung cancer (25), indicating that the statistically significant change scores may be clinically meaningful.

**Figure 2.**
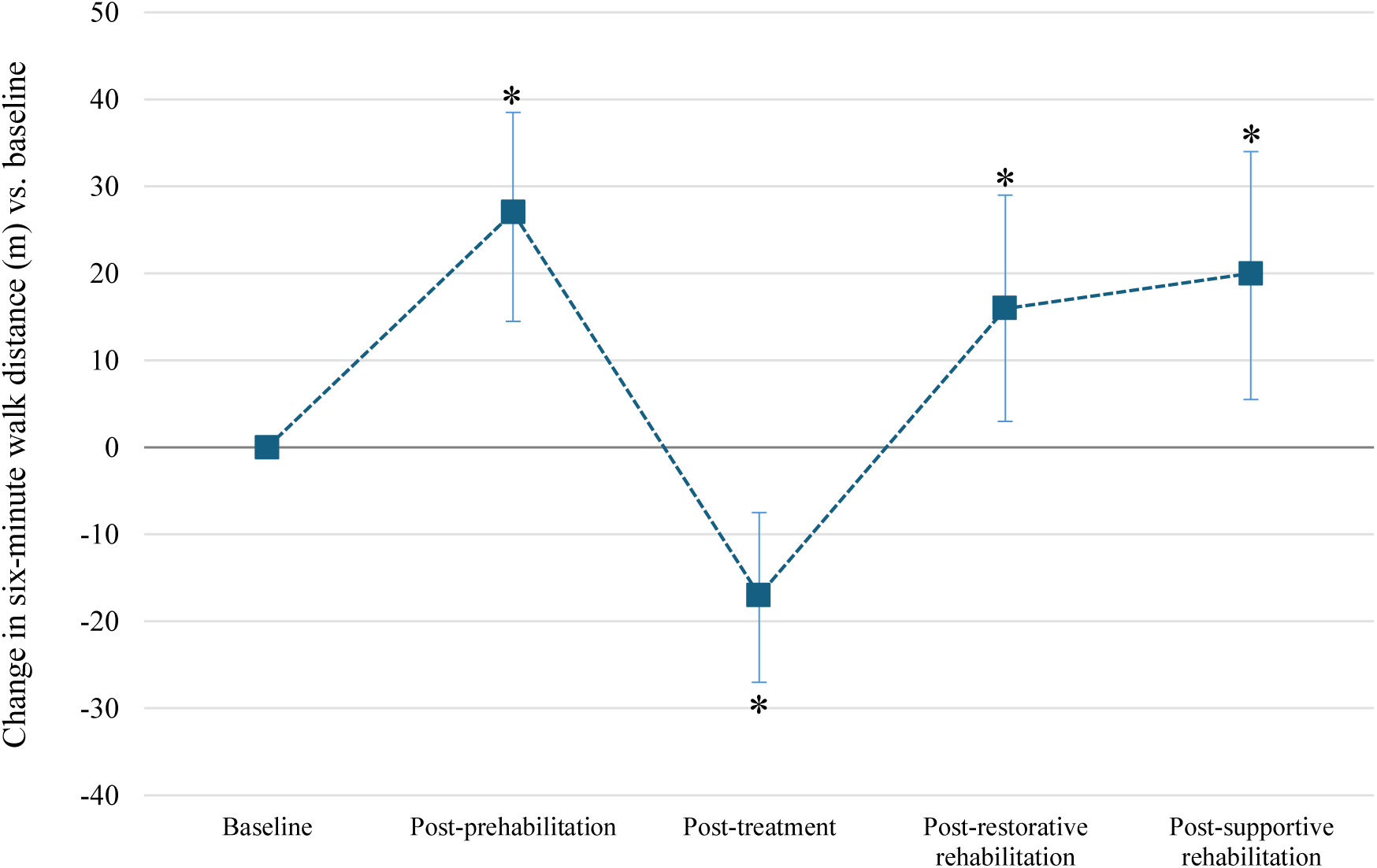
Median change in six-minute walk distance (m) versus baseline (all patients), with 95% confidence intervals. *Indicates a significant difference from baseline.

#### Lower-Limb Strength and Strength-Endurance

##### Sit-to-Stand Performance

The median number of sit-to-stand (STS) repetitions in 30 seconds at baseline was 14.5 for males and 12 for females. For the 60-s STS test, the median baseline scores were 28.5 for males and 23 for females. Compared to normative data, the 30-s STS test scores indicate similar fitness levels to community dwelling elderly adults (aged 60-80 years) (26), yet the 60-s STS test scores are within the lowest quartile (lowest function) for male and female adults aged under 75 years (27), suggesting low strength-endurance. Figure 3 displays changes in both 30-s and 60-s STS performance. At a group level (all patients combined), the median number of sit-to-stand repetitions (reps) increased after prehabilitation by 2 reps for the 30-s and 60-s STS tests. However, only the 30-s STS change score was significantly different from baseline (p<0.01, Cohen’s d=0.9). Sit-to-stand repetitions were significantly lower than baseline in both STS tests after treatment (30s: -1 rep, p=0.03, Cohen’s d=0.48; 60s: -3 reps, p<0.01, Cohen’s d=0.71). Following rehabilitation, 30-s STS ability was restored to baseline, whereas 60-s STS ability was 2.5 reps above baseline (p=0.06, Cohen’s d=0.78), and was 5.5 reps above the post-treatment score but this was not statistically significant (p=0.35, Cohen’s d=0.58). Based on MCID values of 2-3 reps from pulmonary disease populations (28–30), the post-treatment increase in 60-s STS reps may be clinically meaningful.

**Figure 3.**
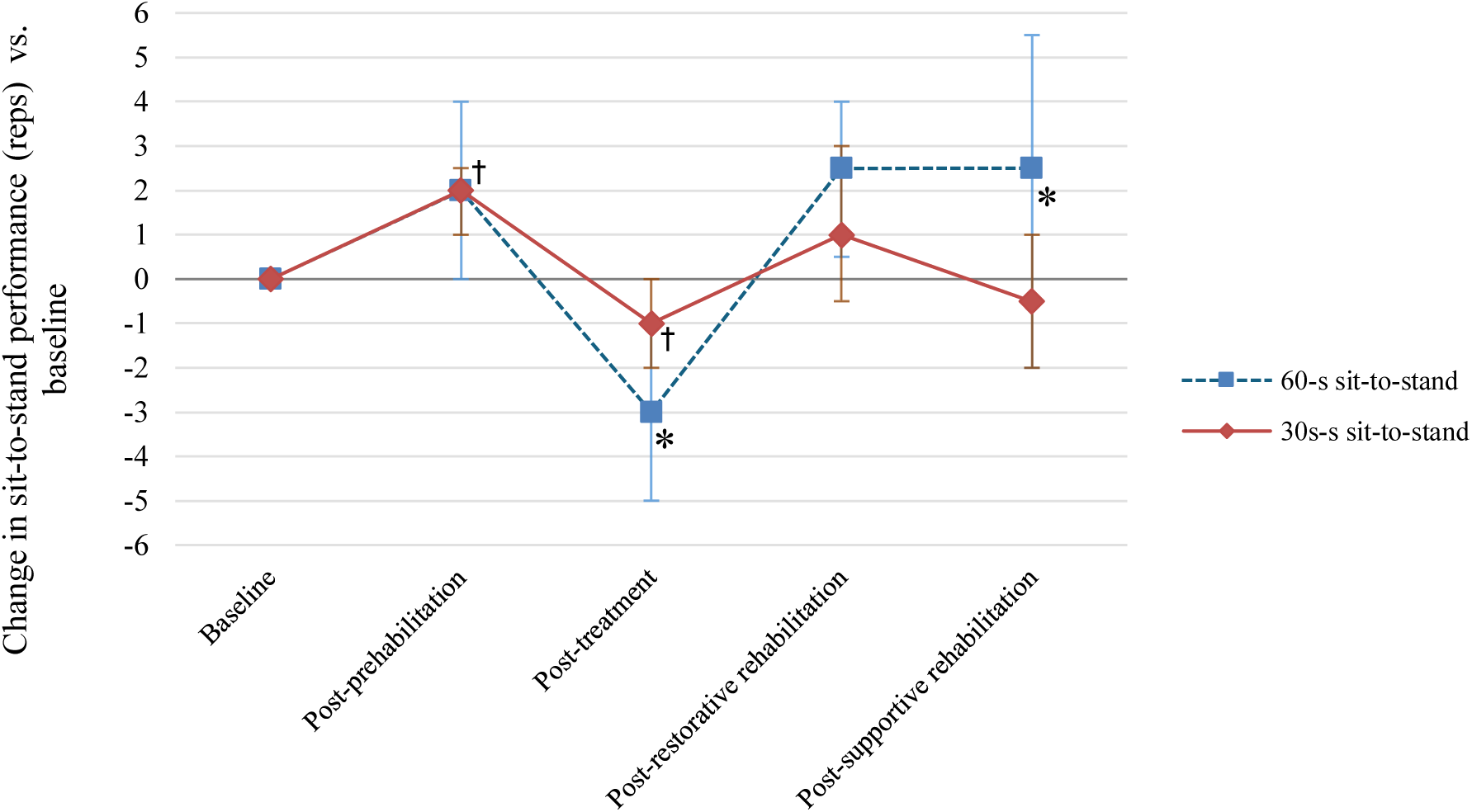
Median change in 30-second and 60-second sit-to-stand repetitions versus baseline (all patients), with 95% confidence intervals. † Indicates a significant difference from baseline for 30-s sit-to-stand; *Indicates a significant difference from baseline for 60-s sit-to-stand.

Figure 4 displays changes in 60-s STS performance categorised by sex. Between-sex differences were identified at the post-treatment assessment (p=0.003), whereby 60-s STS performance worsened in males (−5.5 reps versus baseline) but was slightly above baseline for females (+1.0 rep). By the post-restorative and post-supportive rehabilitation assessments, both 30-s and 60-s STS performance returned to baseline levels, indicating a full recovery of lower-limb function. However, it is noteworthy that females exhibited increased 60-s STS performance at the post-restorative rehabilitation assessment (+4.0 reps), compared to a negligible change in performance for males (+0.1 rep), indicating enhanced recovery in females. The MCID values of 2-3 reps derived from pulmonary disease cohorts (28–30) suggests that the 2-rep increase after prehabilitation (all groups combined), the 5.5-rep decline in males post-treatment and subsequent recovery to baseline, and the 4-rep improvement in females post-restorative rehabilitation are all likely to be clinically meaningful.

**Figure 4.**
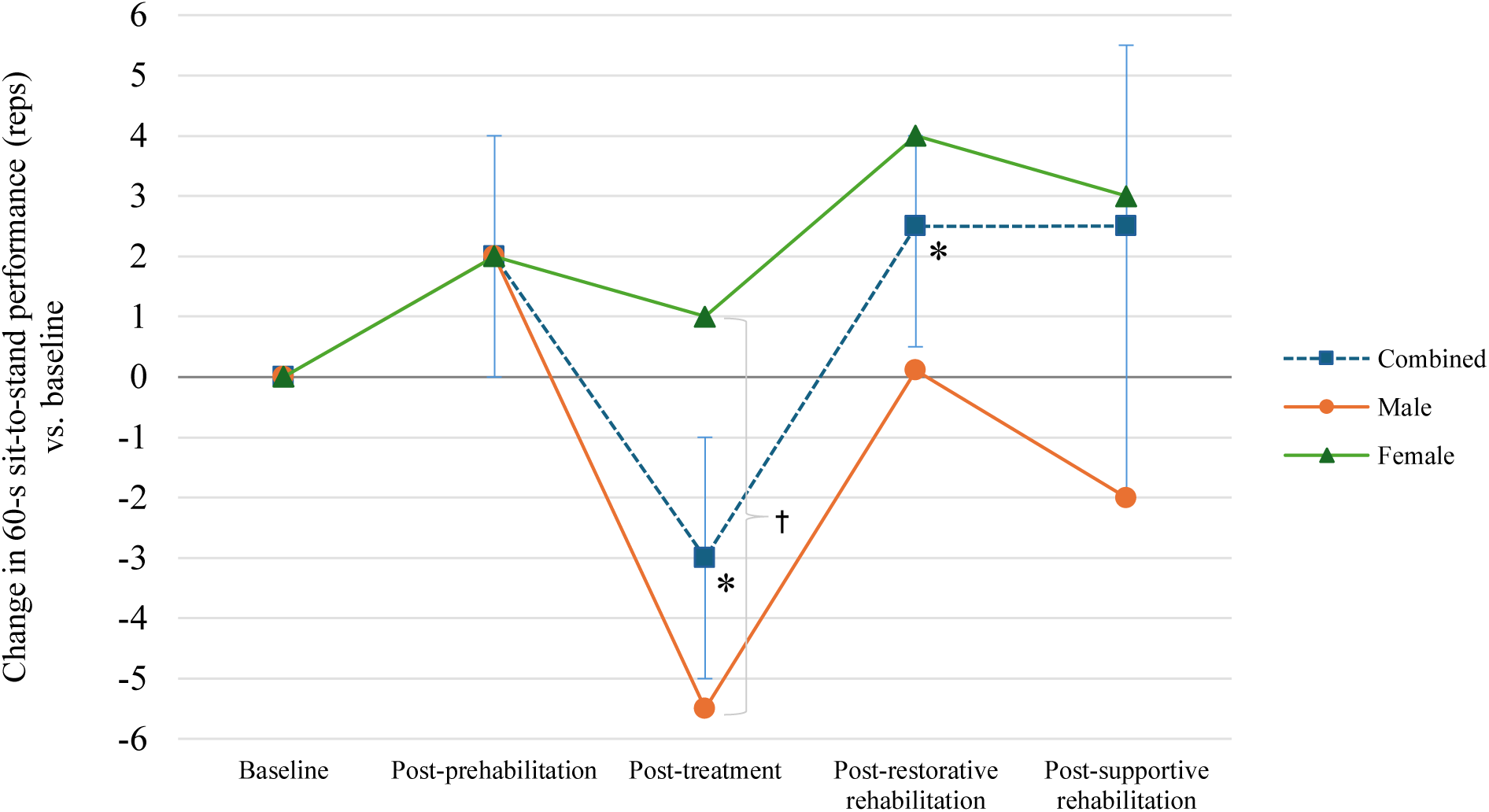
Median change in 60-second sit-to-stand repetitions versus baseline. *Indicates a significant difference from baseline for the combined cohort (all patients); † indicates a significant between group difference. 95% confidence intervals are displayed for the combined cohort only.

##### Upper-Limb Strength

At baseline, the median handgrip strength for females was 20kg and 19kg for the dominant and non-dominant hand, respectively. In addition, the median baseline handgrip strength for males was 33kg and 32kg for the dominant and non-dominant hand, respectively. Both sexes’ handgrip strength scores are in the lowest 10^th^ percentile for both sexes in the 65-69 age category (46) and indicate frailty (47). Figure 5 displays changes in non-dominant handgrip strength for all patients combined, and categorised by sex. At a group level (all patients combined), dominant and non-dominant handgrip strength did not significantly change after prehabilitation (both hands: p>0.47, Cohen’s d<0.15), then significantly decreased (both hands: ≈-2kg, p<0.01, Cohen’s d>0.99) following treatment, and remained lower than baseline levels. There were statistically significant between-sex differences observed for non-dominant handgrip strength at the post-restorative (male: -2.7kg; female -0.5kg, p<0.05) and post-supportive (male:-2.1kg; female +0.6kg, p<0.05) rehabilitation assessments, with men exhibiting a greater decline compared to baseline, relative to females. This suggests that females’ upper limb strength recovered to a greater extent following cancer treatment, whereas males experienced a sustained decline in upper limb strength.

**Figure 5.**
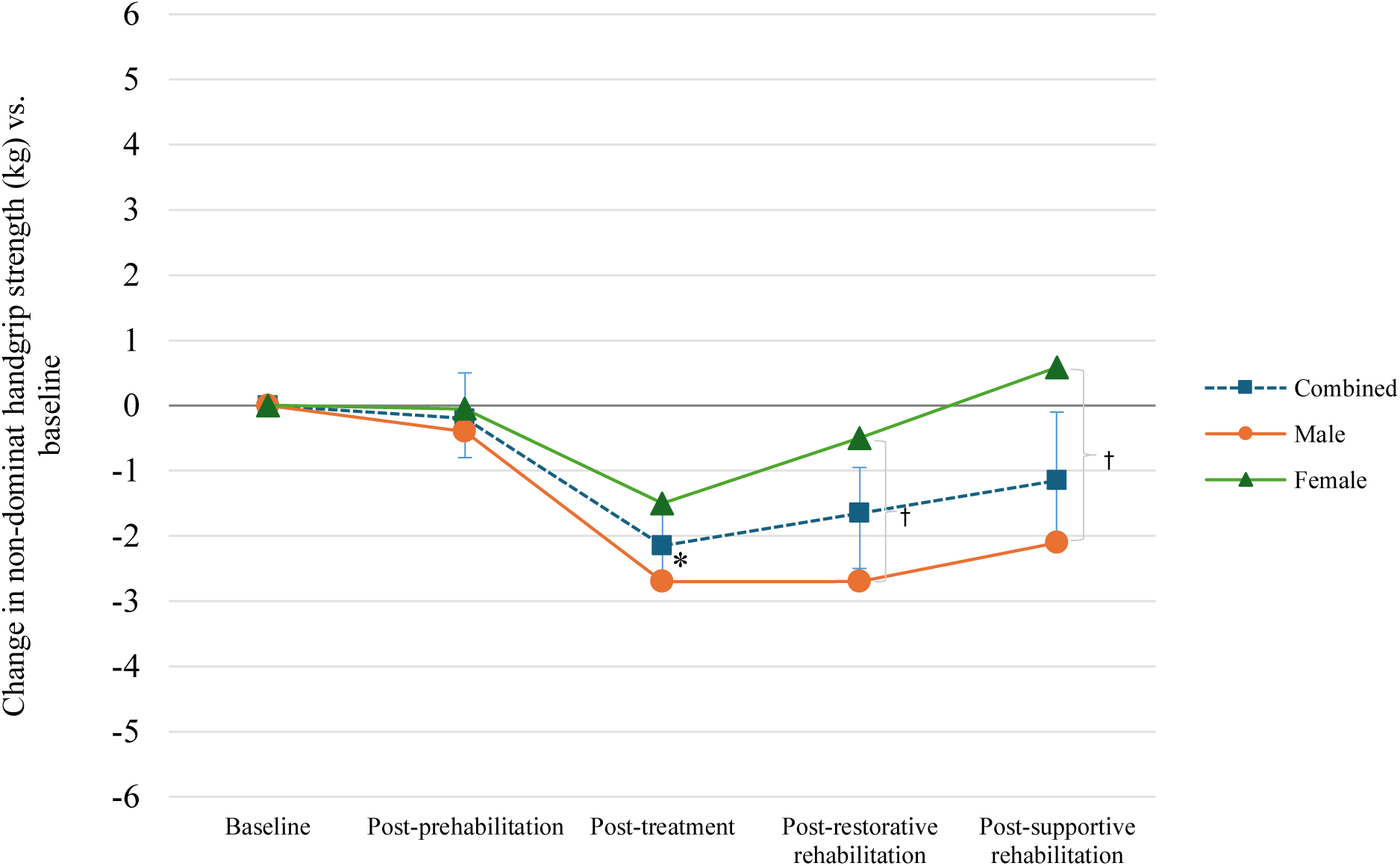
Median change in non-dominant handgrip strength (kg) versus baseline. *Indicates a significant difference from baseline for the combined cohort (all patients); † indicates a significant between-group difference. 95% confidence intervals are displayed for the combined cohort only.

Upper GI cancer patients exhibited a significantly larger post-treatment decline in dominant (−3.7 kg) and non-dominant handgrip strength (−4.3 kg) compared to colorectal (dominant hand: −1.5 kg; non-dominant hand: −1.35 kg; p<0.05) and lung cancer cohorts (dominant hand: -1.1 kg; dominant hand:-0.7 kg; p<0.05), relative to baseline (all p<0.05) (Figure 6). While the MCID for handgrip strength in cancer patients has not been established, a range of 5-6.5 kg is recommended for older adults (31). Except for the post-treatment decline in handgrip strength in the upper GI cohort, which approached the MCID, none of the changes in handgrip strength in the other tumour groups exceeded this threshold.

**Figure 6.**
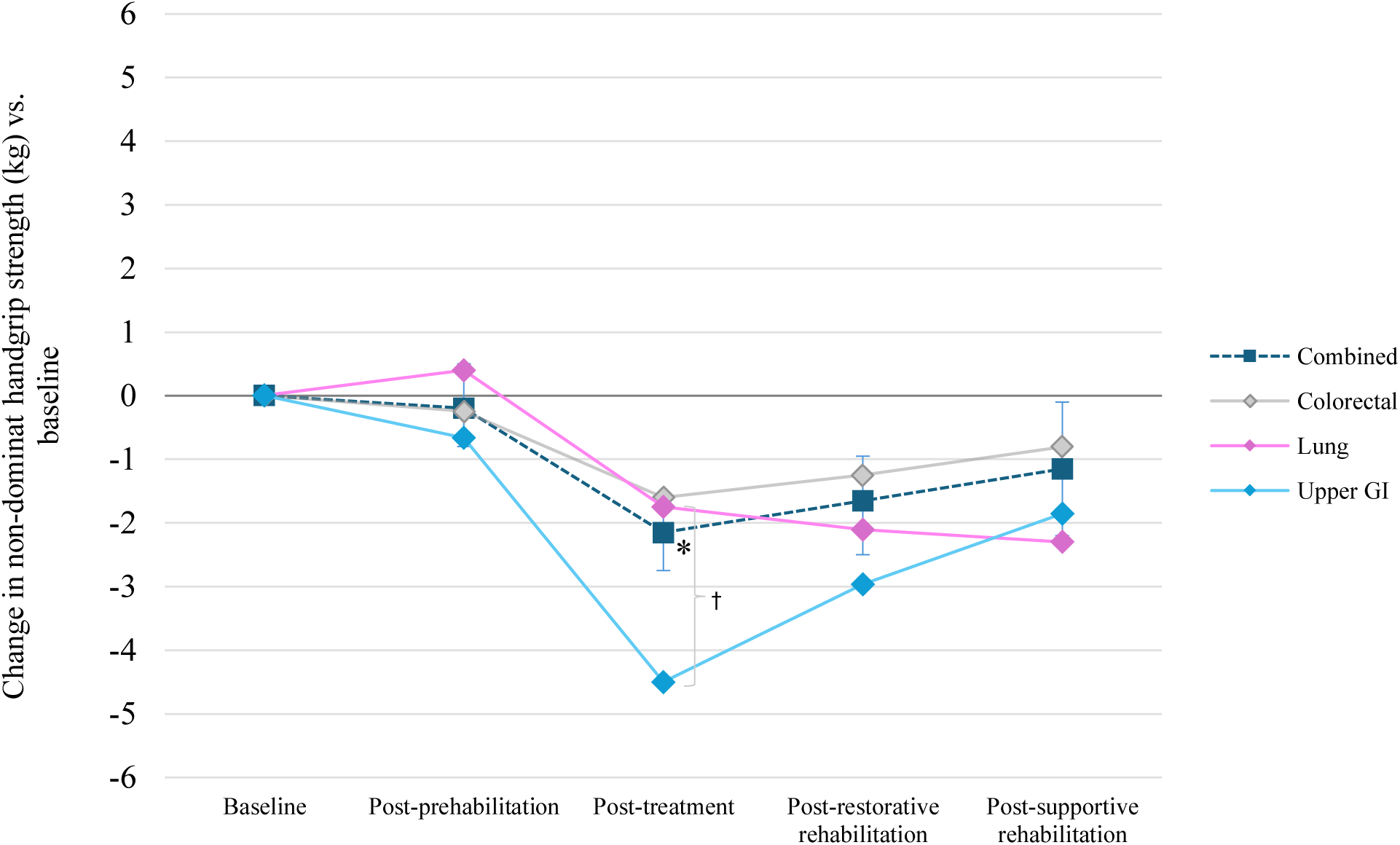
Median change in non-dominant handgrip strength (kg) versus baseline. *Indicates a significant difference from baseline for the combined cohort (all patients); † indicates a significant between group difference. 95% confidence intervals are displayed for the combined cohort only.

### Fatigue

The FACIT-Fatigue Scale ranges from 0 to 52, with lower scores indicating higher fatigue (worse outcomes). The median baseline FACIT score was 39 for male and 38 for female patients, which is lower than the general population with normative scores typically ranging from 43 to 46 for adults aged 50 to 70 years (32). Figure 7 displays changes in fatigue levels for all patient cohorts combined; no between-group differences were identified. Fatigue improved after prehabilitation (+2.5 points, p<0.01), worsened following treatment (−3.5 points compared to baseline, p<0.01) and subsequently returned to pre-treatment levels after the restorative and supportive rehabilitation phases (p>0.05, not different to baseline). Two studies have reported MCIDs for FACIT-Fatigue in cancer populations, ranging widely from 3 to 17 points (32,33). Viewed in the context of MCIDs, the relatively small group-level changes in fatigue may not be clinically relevant, apart from the expected worsening of fatigue measured at the post-treatment assessment (−3.5 points).

**Figure 7.**
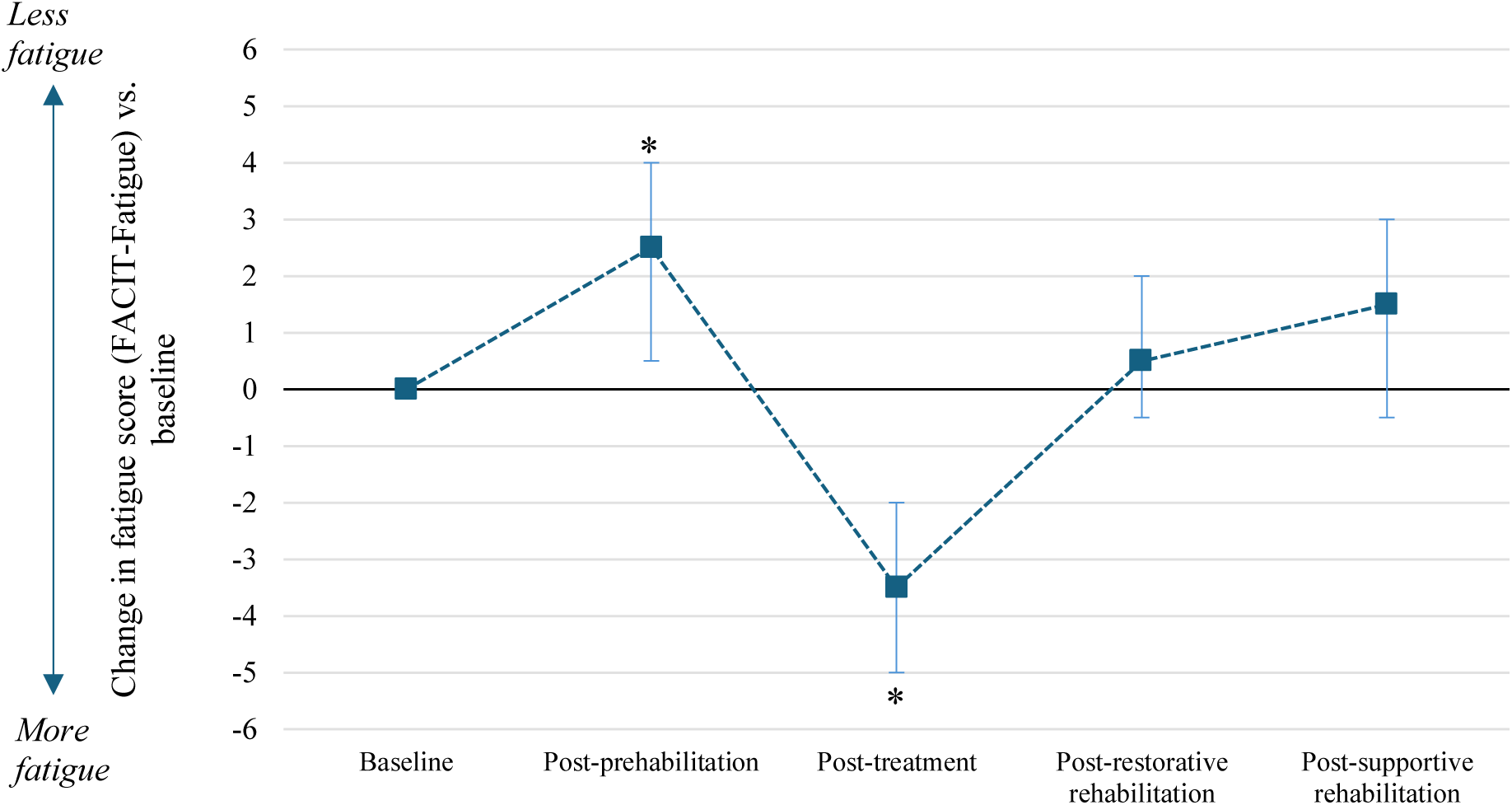
Median change in fatigue score (FACIT-Fatigue) versus baseline (all patients), with 95% confidence intervals. *Indicates a significant difference from baseline.

### Psychological Wellbeing Outcomes

#### Quality of Life

The EQ-5D-5L index scale ranges from -0.594 (worst possible health state) to 1 (best possible health state), whereas the EQ-5D visual analogue scale (VAS) ranges from 0 (worst imaginable health state) to 100 (best imaginable health state). Baseline median scores were 0.76 for the EQ-5D-5L index and 65 for the VAS. The baseline median index score aligns with normative values for adults aged 50–80 years (0.73–0.80 points), but the VAS score is notably lower than the normative range (74–82 points) for this age bracket. There were no between-sex or between-cancer type differences in quality of life; therefore, pooled data are presented. Figures 8 and 9 display changes in EQ-5D-5L index and VAS scores, respectively. With regards to index scores, quality of life significantly declined by a median of 0.06 points (p<0.05) following cancer treatment and returned to baseline after the subsequent rehabilitation phases (p>0.05). No other significant differences in EQ-5D-5L index scores were observed. In contrast, the median EQ-5D VAS score significantly improved after prehabilitation (+7.5 points, p<0.01), returned to baseline after treatment (p>0.05), and significantly improved beyond baseline after both the restorative (+10 points, p<0.01) and supportive rehabilitation phases (+7.5 points, p<0.01) indicating a sustained improvement in quality of life after cancer treatment. For context, the MCID for the EQ-5D-5L in cancer cohorts is estimated to be 0.06–0.08 for the index score and 7–10 points for the VAS score (34). This suggests that the changes in VAS scores are clinically meaningful.

**Figure 8.**
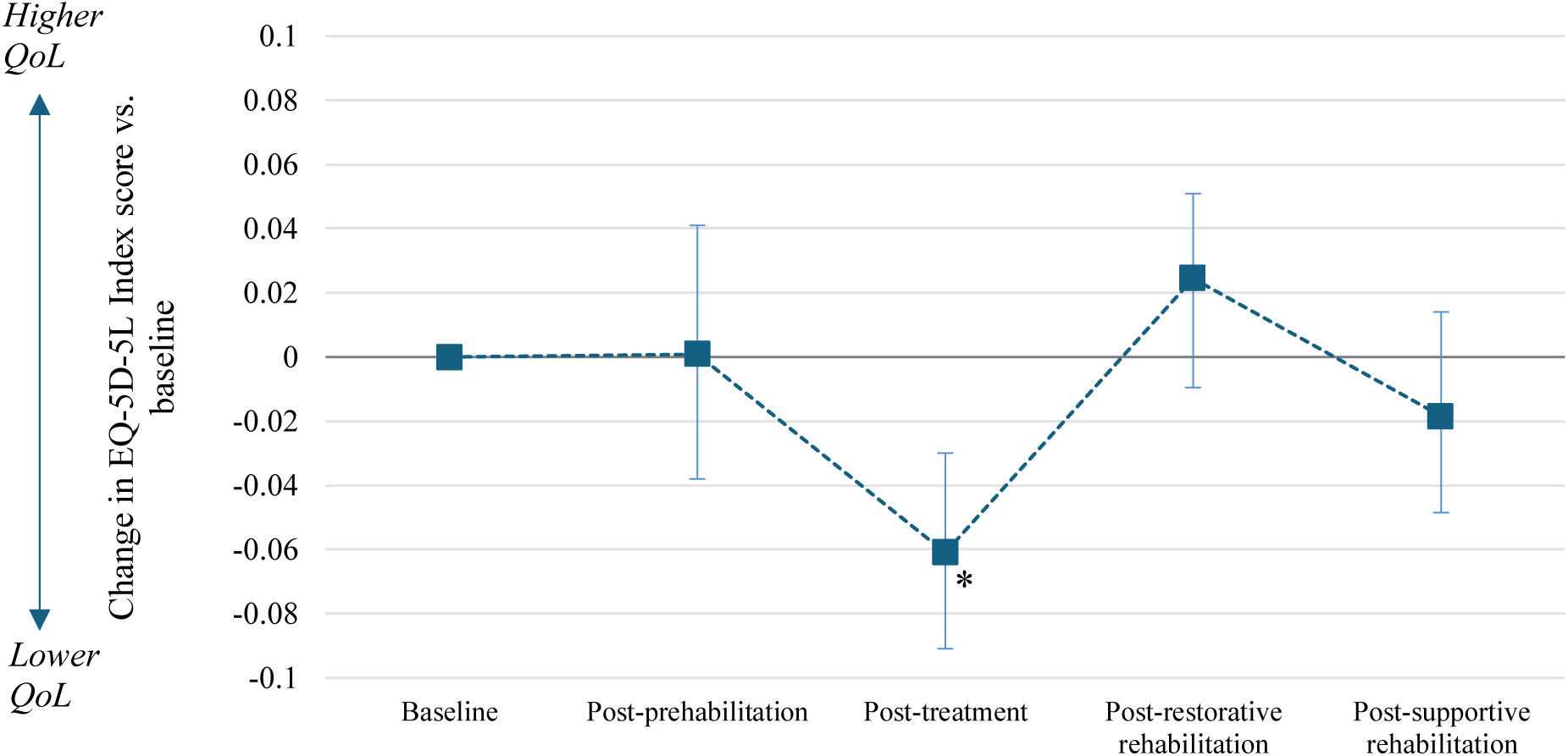
Median change in EQ-5D-5L Index score versus baseline (all patients), with 95% confidence intervals. *Indicates a significant difference from baseline. QoL, quality of life

**Figure 9.**
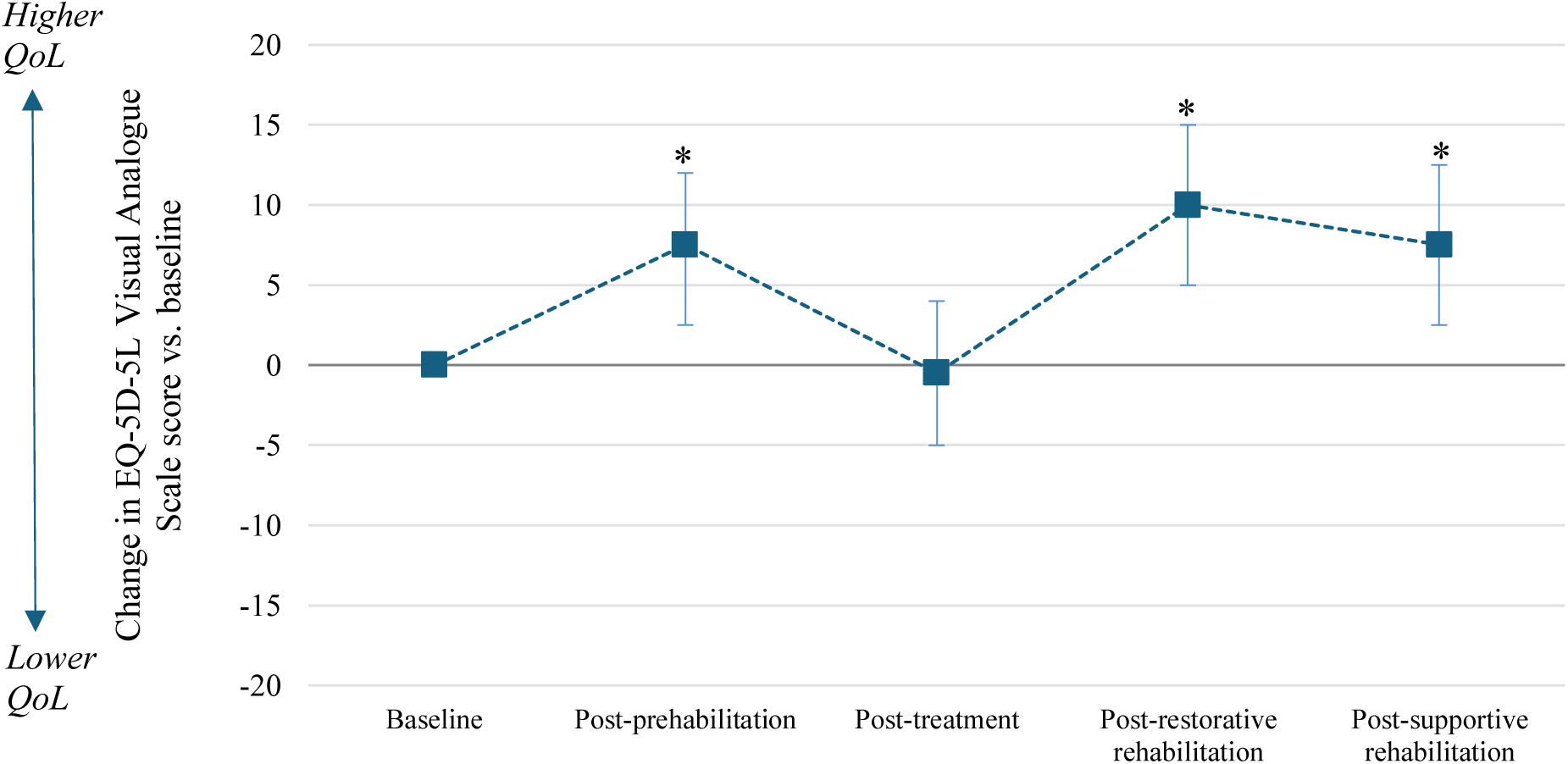
Median change in EQ-5D-5L Visual Analogue Scale score versus baseline (all patients), with 95% confidence intervals. *Indicates a significant difference from baseline.

#### Anxiety

The 7-item Generalized Anxiety Disorder (GAD-7) scale ranges from 0 to 21, with higher scores representing more severe anxiety symptoms. The median baseline GAD-7 score was 5.5, which lies between normative values for the general population (3±3; mild anxiety, (35)) and patients with cancer-related pain and or depression (8±6; mild to moderate anxiety, (18)). There were no between-group differences in anxiety symptoms, therefore pooled data are presented. Throughout the Active Together pathway, anxiety symptoms progressively declined (improved), reaching -2 points after the restorative phase which was sustained throughout the supportive phase (all p< 0.01). Figure 10 displays changes in anxiety symptoms for all patient cohorts combined. The MCID for the GAD-7 is not established for cancer patients, however, a 4-point change has been recommended as a clinically meaningful difference in patients with chronic depression (36).

**Figure 10.**
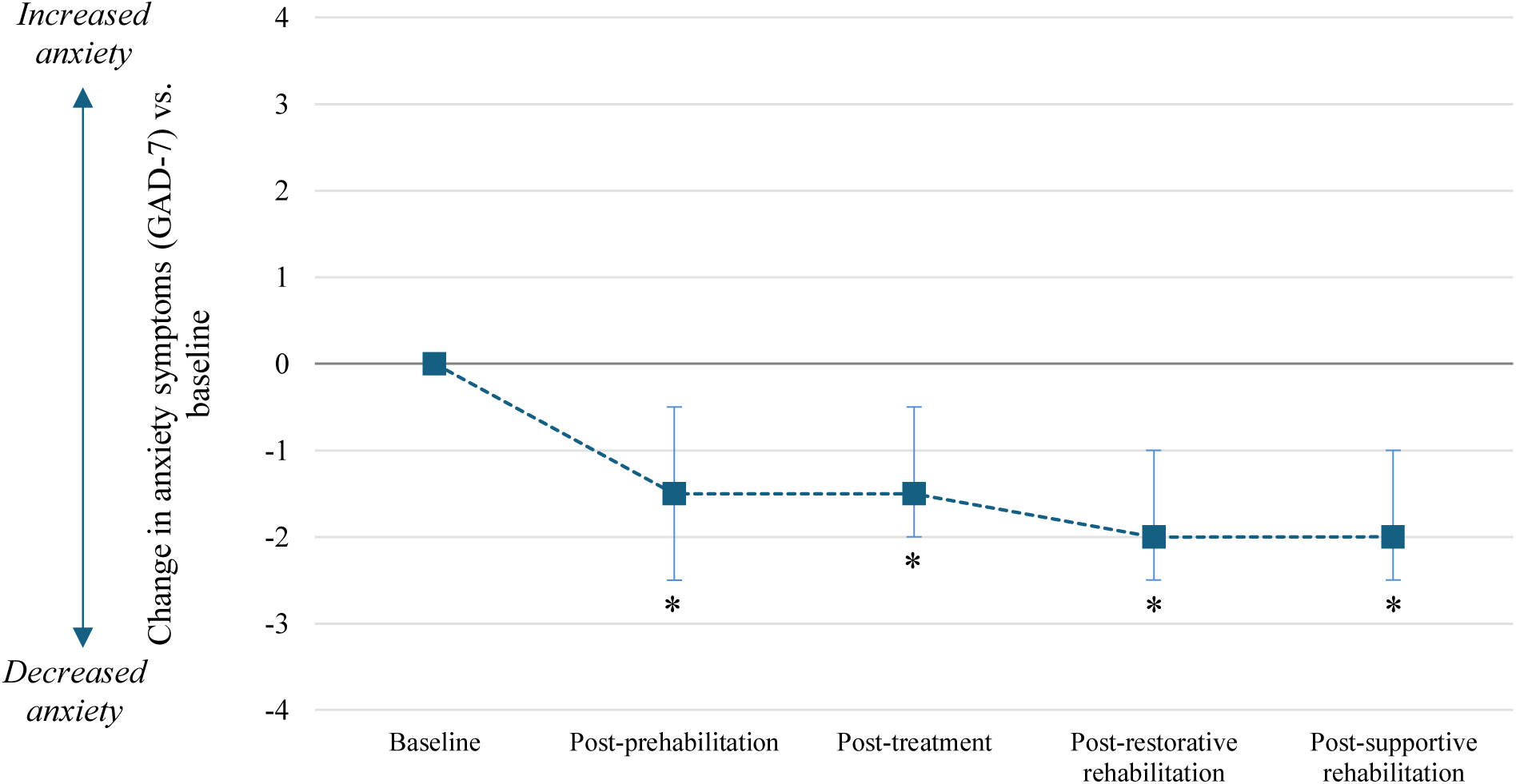
Median change in anxiety symptoms (GAD-7) versus baseline (all patients), with 95% confidence intervals. *Indicates a significant difference from baseline.

#### Depression

The 9-item Patient Health Questionnaire-9 (PHQ-9) assesses depression symptoms on a scale of 0 to 27, with higher scores indicating greater depression severity. A normative PHQ-9 score for various cancer populations is approximately 5 points (derived from 15 cancer types), ranging between 4 (prostate cancer) and 8 (thyroid cancer) (37). The median baseline PHQ-9 score was 6 (mild depression), which is higher (more depressed) than the general population (normative range: 3-4.5 points) and similar to other cancer populations. After prehabilitation, the median PHQ-9 score reduced (improved) by 1 point below baseline (p<0.01), then returned to baseline after treatment (p>0.05 compared to baseline), and subsequently decreased (improved) by 1.5 points below baseline after restorative rehabilitation (p<0.001) which was sustained through the supportive phase (p=0.01).

There were significant differences between cancer cohorts in depression symptoms at the post-treatment assessment. Compared to colorectal cancer patients who had a median reduction in depression symptoms (−1 point from baseline), lung and upper GI cancer cohorts both had increased depression symptom scores (both differences p<0.05, +2 points from baseline). The MCID for PHQ-9 in patients with cancer-related pain and or depression is estimated to be 2–6 points (18). Except for the post-treatment increase in depression symptoms in lung and upper GI cancer patients, most statistically significant changes in depression symptoms were smaller than established MCIDs. Figure 11 displays changes in depression symptoms for all patient groups combined, and each tumour group.

**Figure 11.**
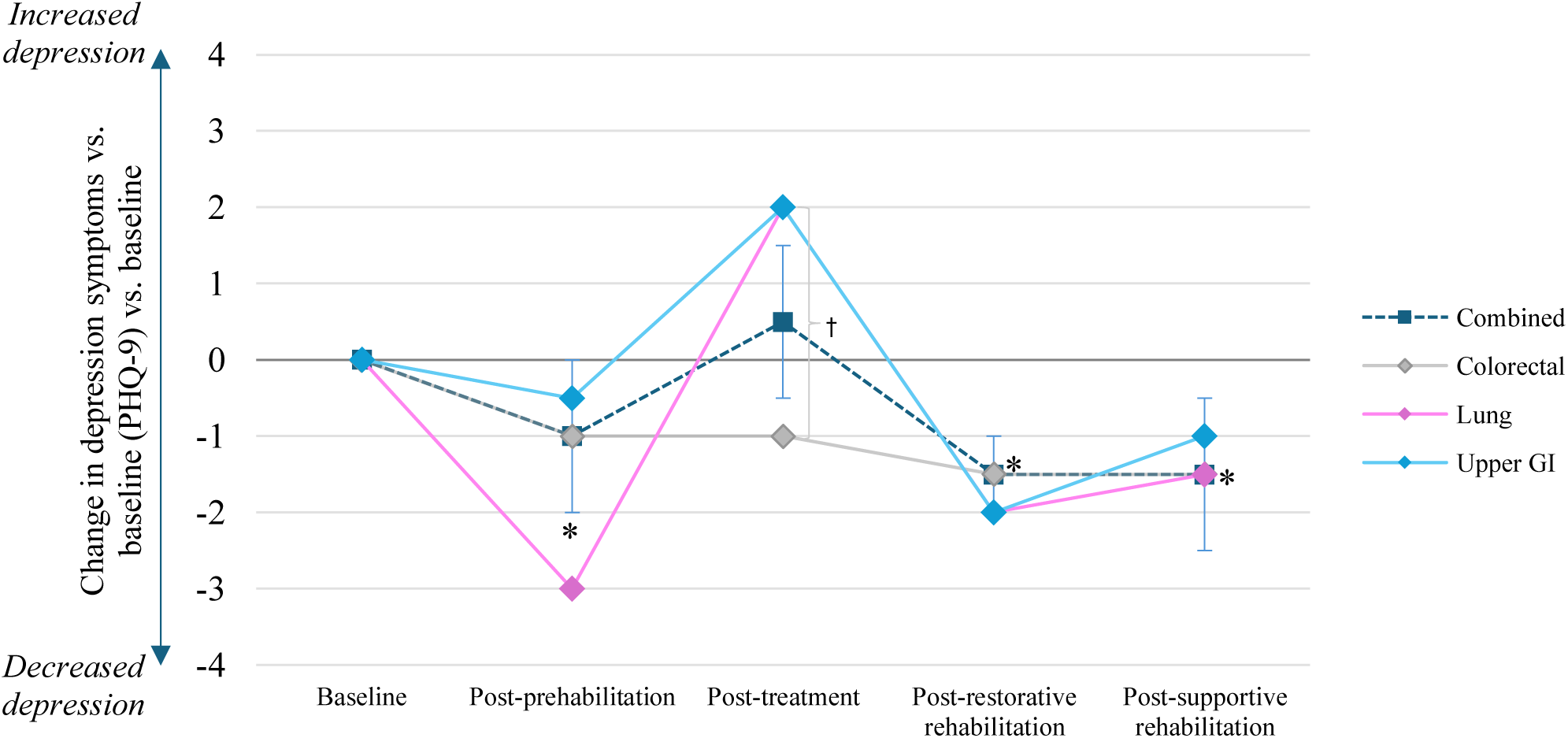
Median change in depression symptoms (PHQ-9) versus baseline.*Indicates a significant difference from baseline for the combined cohort (all patients); † indicates a significant between group difference. 95% confidence intervals are displayed for the combined cohort only.

## Discussion

This clinician-led cancer prehabilitation and rehabilitation programme achieved high uptake (80.9% acceptance rate) and engagement, with 93% completing the prehabilitation phase and 62% completing the full programme over a median duration of 44 weeks. These engagement rates compare favourably to similar clinical rehabilitation programmes (38–40). For example, Manchester’s Prehab4Cancer service reported a 48% prehabilitation completion rate in a lung cancer patient cohort (41), and a prehabilitation completion rate of 73% in a combined cohort of patients with colorectal, lung and upper GI cancer (42).

Patients experienced clinically meaningful improvements in cardiorespiratory fitness (6MWT: +27m post-prehabilitation, +20m post-rehabilitation) and quality of life (EQ-5D VAS: +7.5–10 points post-rehabilitation). These results suggest that prehabilitation may increase resilience to treatment, and that rehabilitation may enhance post-treatment recovery, whilst supporting patients’ wellbeing. Notably, the expected post-treatment decline in 6MWT distance (−17 meters) was less severe than in clinical trial control group participants (−28 meters) (23), which may reflect a mitigated loss of post-treatment physical function typically associated with cancer treatment.

Between-sex differences in physical function were observed, with males experiencing a greater, post-treatment decline in 60-second STS performance (−5.5 reps) which was restored to baseline levels after rehabilitation. In the female cohort, 60-s STS function improved above baseline throughout all phases (+2 to +4 reps). Females’ handgrip strength was restored after post-treatment rehabilitation, whereas males’ handgrip strength remained significantly lower than baseline after treatment. Collectively, these between-sex differences highlight that males tend to have a greater decline in physical function after treatment, whereas females have a smaller decline followed by a more complete recovery. These sex differences may be partly explained by surgery type and hormonal factors. Upper GI surgery is generally more invasive than other cancer surgeries and is associated with greater functional impairments (43); notably, 75–78% of the upper GI cohort in this service evaluation were male. Additionally, cancer treatments can cause an abrupt decline in testosterone in males, leading to accelerated muscle atrophy and impaired recovery (44). By contrast, most women in this cohort were post-menopausal and therefore more likely to be living with low levels of both oestrogen and testosterone (45), meaning that cancer treatments may have a less severe impact on muscle mass and function compared to males.

The physical fitness improvements exhibited by Active Together patients following prehabilitation were of a smaller magnitude than those reported by Prehab4Cancer, a comparable service based in Greater Manchester, UK. Specifically, the increase in 6MWT distance was +27 m for Active Together versus +43 m for Prehab4Cancer. Following a similar trend, 60-second STS repetitions increased by +2 reps versus +4.8 reps, for Active Together and Prehab4Cancer patients, respectively. A likely explanation for these differences is exercise intensity; Prehab4Cancer incorporates high-intensity interval training (HIIT), while Active Together exercise sessions were typically moderate intensity. The greater physiological stimulus of HIIT is associated with superior short-term improvements in cardiorespiratory fitness, as demonstrated in UK cardiac rehabilitation programmes (46). The contrasting exercise training approaches may reflect the differences in patient populations, whereby Active Together accepts all patients regardless of health status and risk profile, whereas Prehab4Cancer report excluding patients with very low cardiorespiratory fitness (V̇O_2_max: <10 mL/kg/min^-1^) due to safety concerns. In both services, changes in handgrip strength following prehabilitation were negligible, which may indicate that the resistance training dose was suboptimal for cancer patients. It is also possible that handgrip dynamometry does not adequately capture upper limb strength, given its restriction to the hand and forearm muscles.

Fatigue levels improved modestly during prehabilitation then worsened significantly following treatment as expected. Fatigue scores subsequently returned to baseline levels during the rehabilitation phases, suggesting that multi-modal rehabilitation supported the self-management of cancer-related fatigue—a commonly reported, debilitating symptom in cancer populations (47,48).

Anxiety symptoms (GAD-7) progressively decreased (improved) across the Active Together pathway, although the clinical relevance of these changes remains uncertain due to a lack of cancer-specific MCID data. Depression symptoms (PHQ-9) followed a similar trend, though changes were typically below MCID thresholds. Subgroup analyses revealed that lung and upper GI cancer patients experienced an increase in depression symptoms post-treatment, in contrast to the colorectal cohort, who exhibited a decrease in depression symptoms. The reasons for this discrepancy are not fully understood but may be linked to the greater invasiveness of lung and upper GI surgeries compared to colorectal surgery, as indicated by longer stays in critical care (unpublished data). This finding highlights that patients undergoing more intensive treatments, such as thoracic or upper abdominal surgeries may require more psychological support during the post-treatment phase.

Quality of life assessed using the EQ-5D-5L index score and visual analogue scale had divergent results. The EQ-5D-5L index score did not change after prehabilitation, declined post-treatment and later recovered to baseline levels. This lack of change following prehabilitation contrasts with results reported a Prehab4Cancer evaluation (only index scores were reported), which demonstrated a change of +0.04 points in the EQ-5D index score, although this change was below the established MCID (34). For Active Together patients, the EQ5D VAS scores improved significantly during prehabilitation, returned to baseline after treatment, and improved during the restorative and supportive rehabilitation phases. Notably, VAS scores exceeded MCID thresholds, suggesting that patients experienced meaningful improvements in perceived health status as they progressed through the Active Together pathway. The reasons for the divergence between EQ-5D index and VAS scores remain unclear, but may suggest that the VAS is more sensitive to cancer-related health burden.

## Strengths and Limitations

A key strength of this service evaluation is its inclusion of a large and diverse cohort of cancer patients participating in a real-world, clinical service. Key limitations of this service evaluation are that it lacks a control group, which precludes causal inferences. In addition, the absence of comprehensive data on exercise dose, and dietetic/psychological intervention content limits insights into the mechanisms driving patient outcomes. The current article did not report body composition measures for brevity. Although body mass and stature were routinely recorded as part of clinical care, body mass index is considered poor indicator of body composition as it fails to identify conditions such as sarcopenic obesity (49) — a common consequence of cancer treatment (50). Future service evaluations would benefit from a more accurate measure of body composition, at least in a sub-sample of patients.

The discussed results have been contextualised against MCIDs which has limitations. Where possible, MCIDs have been sourced from similar populations, for example, older adults with cancer. However, without considering the phase of cancer treatment, MCIDs could understate the clinical significance of outcomes such as the post-treatment restoration of physical function and psychological wellbeing, and mitigation in post-treatment deterioration. For instance, in people undergoing highly invasive surgery, returning to pre-treatment physical fitness is undoubtedly a clinically meaningful outcome. Yet, such improvements may not be fully captured when benchmarked solely against MCIDs. Further research is needed to establish MCIDs for a wider range of cancer types across the different phases of the cancer pathway.

As a counterpoint, it is important to recognise that cancer patients could return to their pre-treatment state without being offered structured rehabilitation. For example, in the GOSAFE study, which tracked recovery trajectories following major colorectal surgery, 78.6% and 70.6% of the colon and rectal cancer patients, respectively, recovered to their pre-operative functional status three months after treatment without formal prehabilitation or rehabilitation (51). However, this study also highlighted that people with poorer baseline physical function and a higher number of comorbidities were more likely to experience poorer treatment outcomes. A key limitation of the current service evaluation is the absence of comorbidity data and the lack of subߞanalyses stratified by comorbidity status and baseline physical fitness—two important prognostic factors. Future service evaluations should aim to account for these covariates to better understand differential responses to the intervention.

## Conclusion

This service evaluation demonstrates that the Active Together multi-modal cancer prehabilitation and rehabilitation service supports patients with colorectal, lung, and upper GI cancers in preparing for and recovering from cancer treatments. Clinically meaningful improvements in physical fitness and quality of life were achieved. In addition, most other outcome measures were restored to baseline after rehabilitation, indicating successful recovery from cancer treatment. However, male participants and those with upper GI cancer experienced slower and less complete recovery.

## Next Steps

Active Together is currently being rolled out across Yorkshire, engaging a broader range of cancer populations. The programme is adopting flexible delivery models tailored to diverse geographic and service contexts, spanning both NHS and community-based organisations. Maintaining the integrity of the original service while enabling flexibility for local adaptation will be critical for successful expansion. Future work will include a health economics analysis and more detailed information on intervention characteristics and fidelity to understand which elements are most effective and scalable.

## Open Access Statement

For the purpose of open access, the author has applied a Creative Commons Attribution (CC BY) licence to any Author Accepted Manuscript version arising from this submission.

## Data Availability

All data produced in the present study are available upon reasonable request to the authors

## Appendix

**Table 2.**
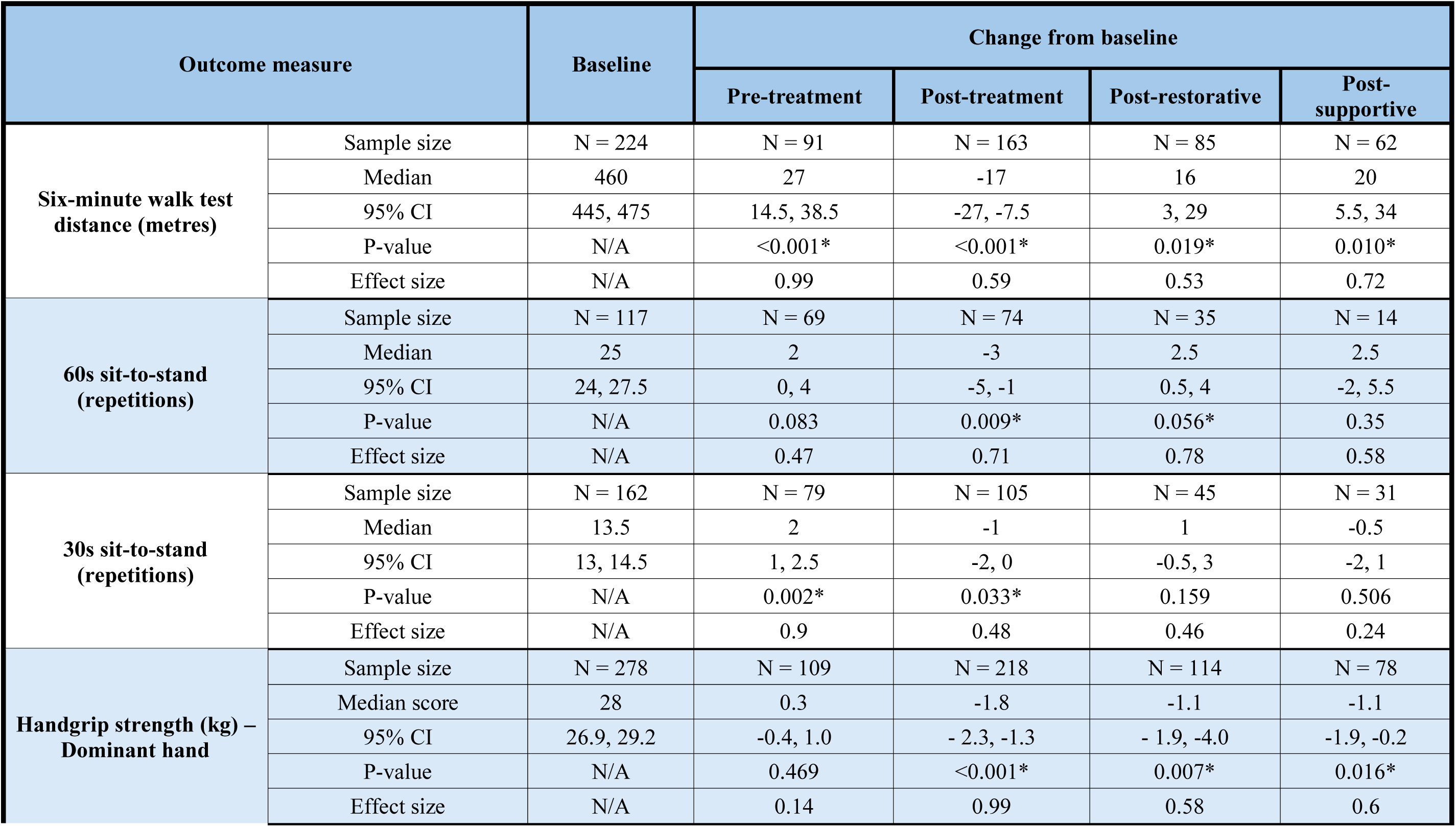
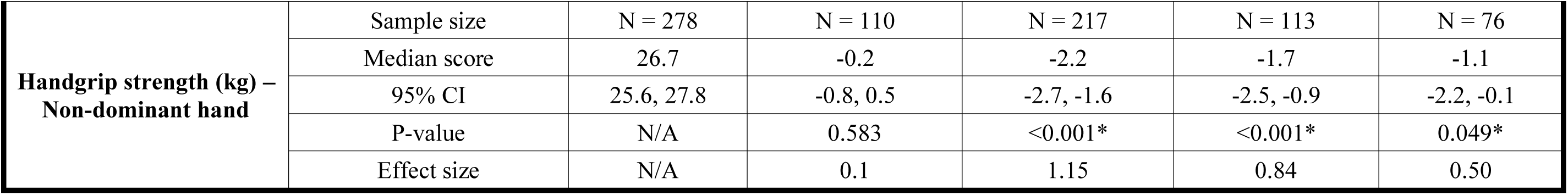
Physical fitness outcome data.

**Table 3.**
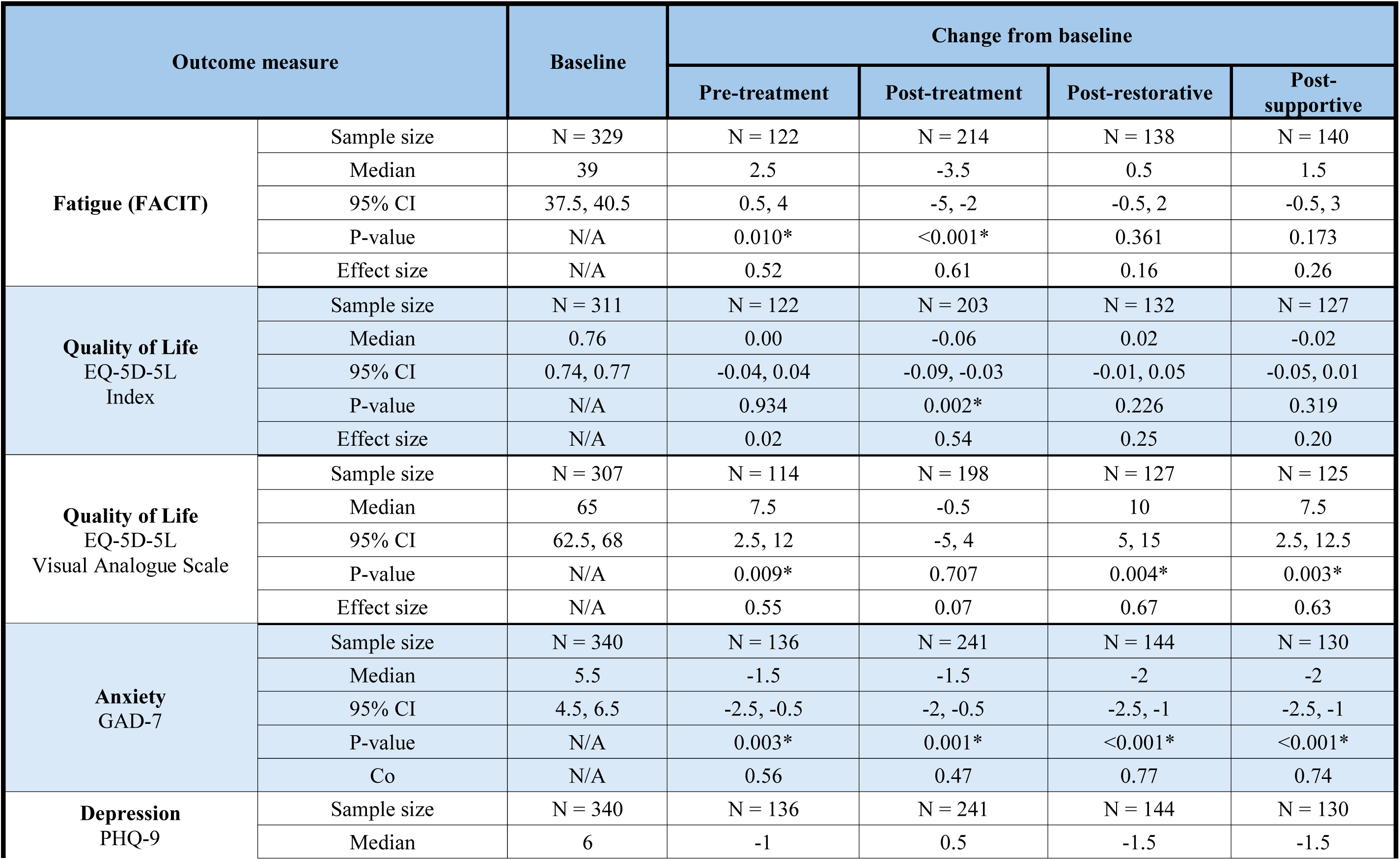
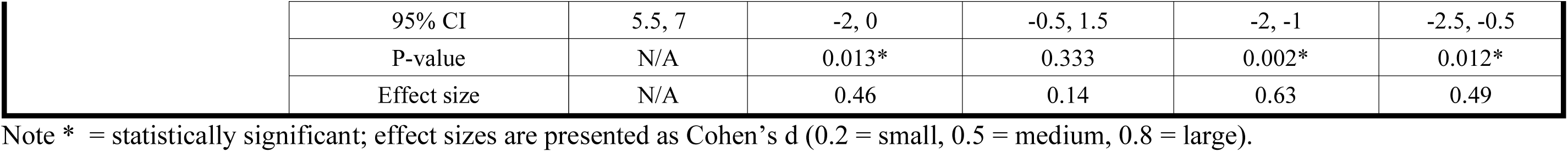
Fatigue and psychological wellbeing outcome data.

